# Understanding Spatial Heterogeneity of COVID-19 Pandemic Using Shape Analysis of Growth Rate Curves

**DOI:** 10.1101/2020.05.25.20112433

**Authors:** Anuj Srivastava, Gerardo Chowell

## Abstract

The growth rates of COVID-19 across different geographical regions (e.g., states in a nation, countries in a continent) follow different shapes and patterns. The overall summaries at coarser spatial scales that are obtained by simply averaging individual curves (across regions) obscure nuanced variability and blurs the spatial heterogeneity at finer spatial scales. We employ statistical methods to analyze shapes of local COVID-19 growth rate curves and statistically group them into distinct clusters, according to their shapes. Using this information, we derive the so-called elastic averages of curves within these clusters, which correspond to the dominant incidence patterns. We apply this methodology to the analysis of the daily incidence trajectory of the COVID-pandemic at two spatial scales: A state-level analysis within the USA and a country-level analysis within Europe during mid-February to mid-May, 2020. Our analyses reveal a few dominant incidence trajectories that characterize transmission dynamics across states in the USA and across countries in Europe. This approach results in broad classifications of spatial areas into different trajectories and adds to the methodological toolkit for guiding public health decision making at different spatial scales.

**Highlights:**

- Coarsely summarizing epidemic data collected at finer spatial scales can result in a loss of heterogenous spatial patterns that exist at finer scales. For instance, the average curves may give the impression that the epidemic’s trajectory is declining when, in fact, the trajectory of the epidemic is increasing in certain areas.
- Shape analysis of COVID-19 growth rate curves discovers significant heterogeneity in epidemic spread patterns across spatial areas which can be statistically clustered into distinct groups.
- At a higher level, clustering spatial patterns into distinct groups helps discern broad trends, such as rapid growth, leveling off, and slow decline in epidemic growth curves resulting from local transmission dynamics. At a finer level, it helps identify temporal patterns of multiple waves that characterize rate curves for different clusters.
- Quantitative methods for characterizing the spatial-temporal dynamics of evolving epidemic emergencies provide an objective framework to understand transmission dynamics for public health decision making.

## 1 Introduction

The ongoing pandemic of novel coronavirus disease (COVID-19) that erupted in China in December 2019 has already generated substantial morbidity and mortality impact around the world during the first few months of transmission [10]. As the novel coronavirus continues its march around the world, the daily trajectory of the epidemic in terms of new cases or deaths represents a key tool for epidemiologist and public health scientists to quantify the reproduction number, assess the evolution of the doubling time, and evaluate the impact of social distancing strategies in different parts of the word [11, 1]. However, overall epidemic curves, obtained by accumulating or averaging curves over large regions, can hide substantial differences in transmission dynamics that exist at finer spatial scales [6]. These nuanced patterns associated with individual regions may be critical to inform the type and intensity of interventions to bring the epidemics under control [11].

There is a need to better understand the spatial variability in the trajectory of the COVID-19 pandemic in different geographic areas around the world [10]. Indeed, the epidemic curves across different continents or countries may display completely different dynamics at a given time. At a higher level, such dynamics include increasing trends, a leveling off, stationary incidence patterns, and decreasing trends. At a finer level, the growth may be characterized by multiple modes depicting multiple waves of the epidemic [2]. Similarly, the epidemic curves at the subnational level within a country display different dynamics over time. Because the type and intensity of public health interventions are expected to vary across space, classifying and summarizing the spatial-temporal dynamics of the novel coronavirus is key for real-time public health decision making.

Quantitative methods for shape analysis of functional data (see e.g., [12]) help investigate the diversity of the spatial-temporal dynamics of transmission of the COVID-19. They provide an objective framework to characterize the spatial-temporal dynamics of the epidemic in different geographic areas within the same country. This methodology is an important tool in a branch of statistics called *Functional Data Analysis*. It has been used to steady stock markets in finance, weather patterns in meteorology, growth rates in biology, and speech data in signal processing. In the context of the COVID-19 pandemic, clustering of curves has been used to analyze travel patterns of migrants in China [3]. We demonstrate how this methodology can be used to generate representative epidemic curves at broader spatial scales (national level) to avoid losing information that results from aggregating local epidemic curves at coarser spatial scales. We apply this methodology to the analysis of the daily incidence trajectory of the COVID-pandemic at two spatial scales: A state-level analysis within the USA and a country-level analysis within Europe during mid-February to mid-May, 2020. Our analyses reveal characteristic trajectories of the pandemic at the state level in the USA and at the country level in Europe, which provide information that may help guide public health decision making at different spatial scales.

## 2 Data

We analyzed daily series of reported COVID-19 cases at two different levels of spatial aggregation: States within the USA and countries in Europe.

- For the USA analysis, we retrieved daily cumulative case count data from the COVID Tracking Project, a volunteer organization dedicated to collecting and publishing data on the spread of COVID-19 in the United States [14]. Data from multiple sources, such as state or district health departments and trusted news reports, are compiled and assessed for data quality to report the *best* available data for each state. Here we use reported daily state - and national-level cumulative case counts from February 27th, 2020 to May 21st, 2020.
- For the country-level analysis in Europe, we retrieved the data from World Health Organization: Coronavirus disease (C0VID-2019) situation reports on May 23, 2020 [9].

## 3 Methodology

We apply quantitative methods to characterize *shapes* of the epidemic rate curves. The rate curves represent the daily count of new cases of reported COVID-19 cases characterizing the trajectory of the epidemic. While epidemic curves have been analyzed statistically in the past [5], focus on their shapes is a relatively new concept. *Why is it interesting to study shapes of these curves?* By focusing on shapes, one is more interested in the numbers and relative heights of peaks and valleys in a curve, rather than their precise locations. For instance, bell-shape curves will be deemed similar even if their peaks are located with small shifts and some height differences, and these bell curves will be considered different from curves that are mostly increasing. (This is different from past work, for example [7], where all epidemic curves are bell shaped and are classified into different classes according to their means and spreads.) Similarly, curves with different up and down patterns will also be considered different irrespective of the locations of their crossover points. Fig. 1 illustrates this idea pictorially. In the left panel we see a number of curves that differ only in heights and horizontal shifts. We consider all these curves to have the exact same shape. In the right panel we see curves with different numbers, locations, and relative heights of the modes. These curves are deemed to have different shapes and one can quantify the shape differences using tools from shape analysis. By comparing the *shapes* of epidemic curves, representing the transmission dynamics in different geographic areas within the same population, we can classify rate curves into groups or clusters that exhibit a similar growth-decay pattern. There are multiple benefits of this approach. This way we can compute national averages that are better reflective of the actual growth patterns of the states. Secondly, and more importantly, it helps label each state in terms of the state of the pandemic. It can also help us discover predominant patterns in epidemic growth, using data across different locations, times, and scales.

**Figure 1:**
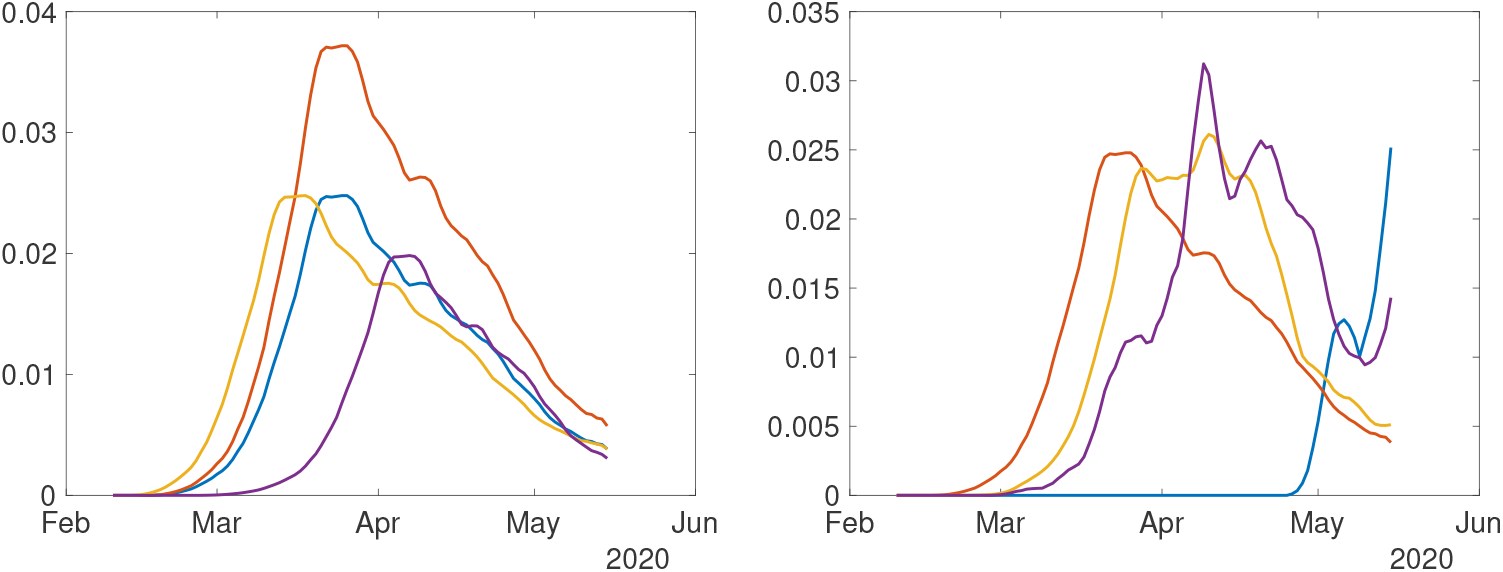
Concept of shapes of curves: All the curves in the left panel are deemed to have the same shape, as they differ only in their heights and horizontal shifts. The curves in the right panel have different shapes.

The next question is: *How does one quantify and statistically analyze shapes of rate curves?* Mathematically, shape is a property that remains unchanged if we rescale axes or translate the curves along axes. In fact, one even allows nonlinear time warping of the time axis, resulting in uneven horizontal shifts of the peaks and valleys, to be a shape-preserving transformation. The invariance of shape to such transformations makes shape analysis a difficult problem. In order to compare and analyze shapes of multiple curves, one has to standardize their domains by scaling axes, normalizing heights, and aligning their peaks and valleys using time warping functions. The resulting curves can then be analyzed for shapes. We employ well-established methodology for shape analysis of functional data described in ref. [12]). This framework provides comprehensive tools for generating statistical summaries and modeling of curves while focusing only on their shapes. Here we apply these tools to trajectory data of the COVID-19 pandemic with the goal of providing sound a framework for sound public health decision making at different spatial scales.

The following sections describe the methodology to analyze and cluster growth rate curves of COVID-19 reported cases. We first pre-process the data to form smooth rate curves for each local unit (a state or a country) over the observation interval. Then, we analyze *shapes* of these rate curves to compare, cluster and summarize growth rates.

### 3.1 Pre-Processing Steps

We start by listing the pre-processing steps applied to COVID-19 daily count data from each unit individually.

- **Time-Differencing**: Since the data includes cumulative counts (or total growth) of positive test counts for different states, we first calculate time differences (approximating time derivatives) of the data to reach *growth rates*. If *f_i_*(*t*) denotes the given cumulative positive counts for state i at time t, then the per-day growth-rate for that state at time t is given by *g_i_(t)* = *f_i_(t)* − *f_i_(t −* 1).
- **Re-Scaling**: The scales of growth rates for some states are very different from other states, due to different population counts, densities, and other variables. In order to separate shape of a curve from its scale, we rescale each curve as follows. We compute the total positive tests for a state, *i.e. r_i_* = ∑*_t_g_i_*(*t*), and then we define *h_i_(t)* = *g_i_(t)/r_i_*.
- **Smoothing**: Next, we smooth these normalized growth rate curves using the **smooth** function in Matlab. With a slight abuse of notation, we shall call the resulting function *h_i_(t*) also. These are the smoothed and normalized growth rate curves, or simply *rate curves* henceforth.

Figure 2 shows an illustration of this processing. The leftmost panel shows the original cumulative positive counts {*f_i_(t*)}, the next panel shows daily new positives {*h_i_(t*)} before smoothing, and the third panel shows rate functions after smoothing and scaling. The rightmost panel shows a coarse overall average of daily positives across states. The top row shows data for US states and the bottom row shows the corresponding data for European countries.

**Figure 2:**
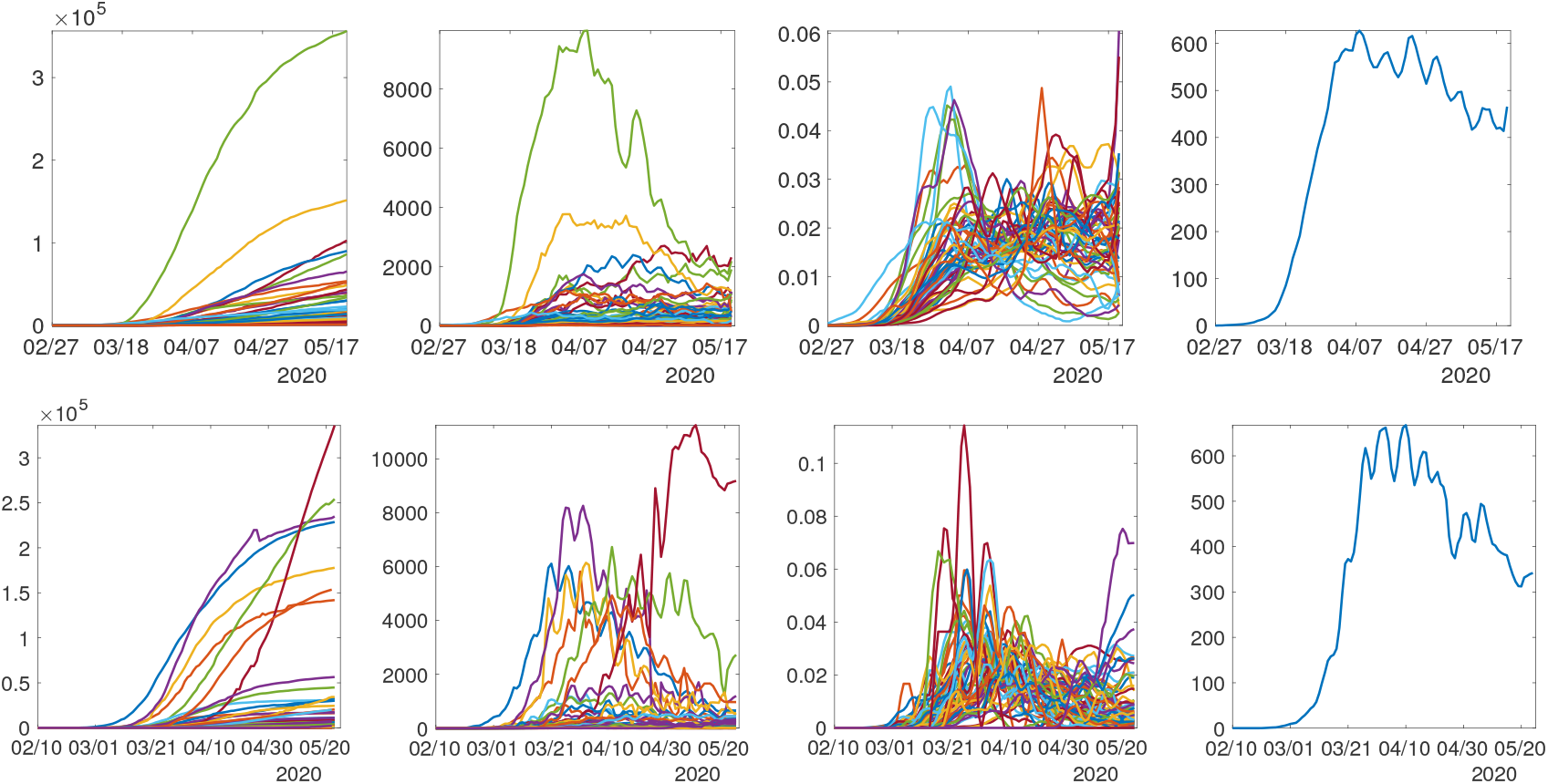
Preprocessing COVID-19 data into growth rate functions. From left to right: Original positive test data; Curves of daily new cases; Smoothed and scaled rate curves; Average of rate curves. Top panels show the state-level data for the USA while the bottom panels display the country-level data for Europe.

### 3.2 Clustering of States

The resulting smoothed and normalized rate curves are then used in statistical analyses. There are several possibilities for this analysis, including modeling, testing, prediction [4, 7] and classification [8]. A raw averaging of data across all states is bound to smooth over interesting patterns and lose interesting smaller structures. The right panels of Fig. 2 show averaging of the daily counts of states for each dataset. Looking at these average curves, one gets the impression that the growth rate is declining universally. However, this conclusion overlooks the variability in dynamics of growth in different states. It is difficult to justify the use of overall averages as representatives of growth patterns. Since our main goal is to extract predominant patterns of growth rates across different states, we start by clustering them into smaller, homogeneous groups. This clustering is important in that it helps recognize spatial heterogeneity of growth rates across geographical regions.

For the purpose of clustering, we use a simple metric to compare any two curves. For any two rate curves, *h_i_* and *h_j_*, we compute the norm ||*h_i_* − *h_j_*||, where the double bars denote the 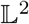 norm of the difference function, i.e. 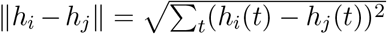. To perform clustering of 51 curves into smaller groups, we apply the dendrogram function in Matlab using the “ward” linkage. The number of clusters is decided empirically based on the display of overall clustering results. We elaborate on this further later on in data analysis.

### 3.3 Alignment and Averaging of Growth Curves

Once we have clustered states into different groups, we seek to derive an appropriate average or a representative curve for each cluster of states. However, a simple arithmetic averaging of curves is not always the best option. In particular, the curves are not perfectly aligned (in terms of their peaks and valleys) and it is well known that direct statistical summaries of misaligned curves often loose structure due to matching of peaks with valleys and vice versa. The solution is to first align peaks and valleys across all curves, using a time-warping algorithm, and then perform averaging. The problem of alignment of curves using time warping is also called *phase-amplitude separation*. In this paper we use the specific alignment algorithm introduced in ref. [13] and described in Chapter 8 of Srivastava and Klassen [12]. For additional details, we refer the reader to the R code package fda_srvf [15].

## 4 Results

The following sections describe the results of our analyses using state-level data for the USA and country-level data for Europe.

### 4.1 State-level analysis for the USA

The normalized rate curves for the period - *2/27/2020 to 05/21/2020* – are shown in the top-right panel of Fig. 2. The results for clustering these 51 curves are shown in Fig. 3. The left side of the figure shows a dendrogram plot - a hierarchical clustering of states. It shows that there are four predominant clusters, which we consider for further analyses. We could also choose five clusters instead, but the results do not change significantly. The resulting four groups are shown in different colors in the dendrogram plot. The right side of the figure shows a color coding of the states - each color represents a different cluster. The listing of states according to this clustering is as follows.

- **Cluster 1:** Iowa, Kansas, Minnesota, Nebraska
- **Cluster 2:** Alabama, Arizona, Arkansas, California, Colorado, Delaware, District of Columbia, Illinois, Indiana, Kentucky, Maryland, Mississippi, New Hampshire, New Mexico, North Carolina, North Dakota, Ohio, Rhode Island, South Dakota, Tennessee, Texas, Utah, Virginia, Wisconsin, Wyoming.
- **Cluster 3:** Connecticut, Florida, Georgia, Maine, Massachusetts, Michigan, Missouri, Nevada, New Jersey, New York, Oklahoma, Oregon, Pennsylvania, South Carolina, Washington, West Virginia.
- **Cluster 4:** Alaska, Hawaii, Idaho, Louisiana, Montana, Vermont

**Figure 3:**
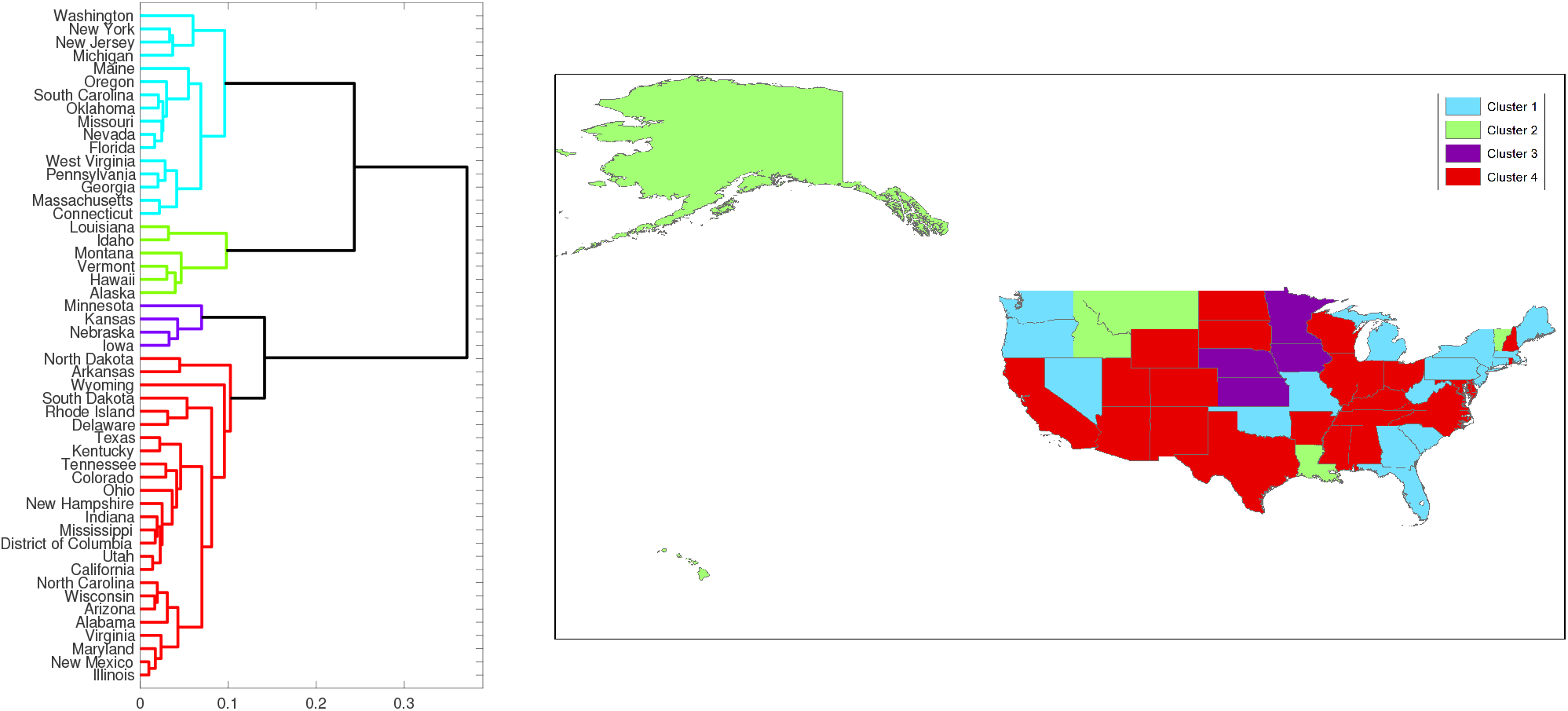
Clustering of US states according to their normalized growth rate curves for the COVID-19 pandemic

The rate curves for these clustered states are shown in Fig. 4, both before (top) and after (bottom) normalization. Since curves for different states have quite different scales, it is not easy to discern overall shape patterns, even within the same cluster in the top row. After rescaling the curves to the same scale, the general trends in the growth rates become clearer. For example, for states in Cluster 4, the rate first goes up sharply and then comes down sharply. For states in Cluster 3, on the other hand, the rate increases sharply at first but has not come down yet, at least not completely.

Although the shapes of curves within a cluster are quite similar, the averaging of these curves still can lose structure. As mentioned earlier, one needs to align the peaks and valleys of curves before averaging. To further extract typical trends, we use an alignment technique from [12, 15] and align rate curves within each cluster. Fig. 5 shows these aligned rate curves in each cluster; the top row shows the unscaled curves while the bottom row shows the scaled curves.

**Figure 4:**
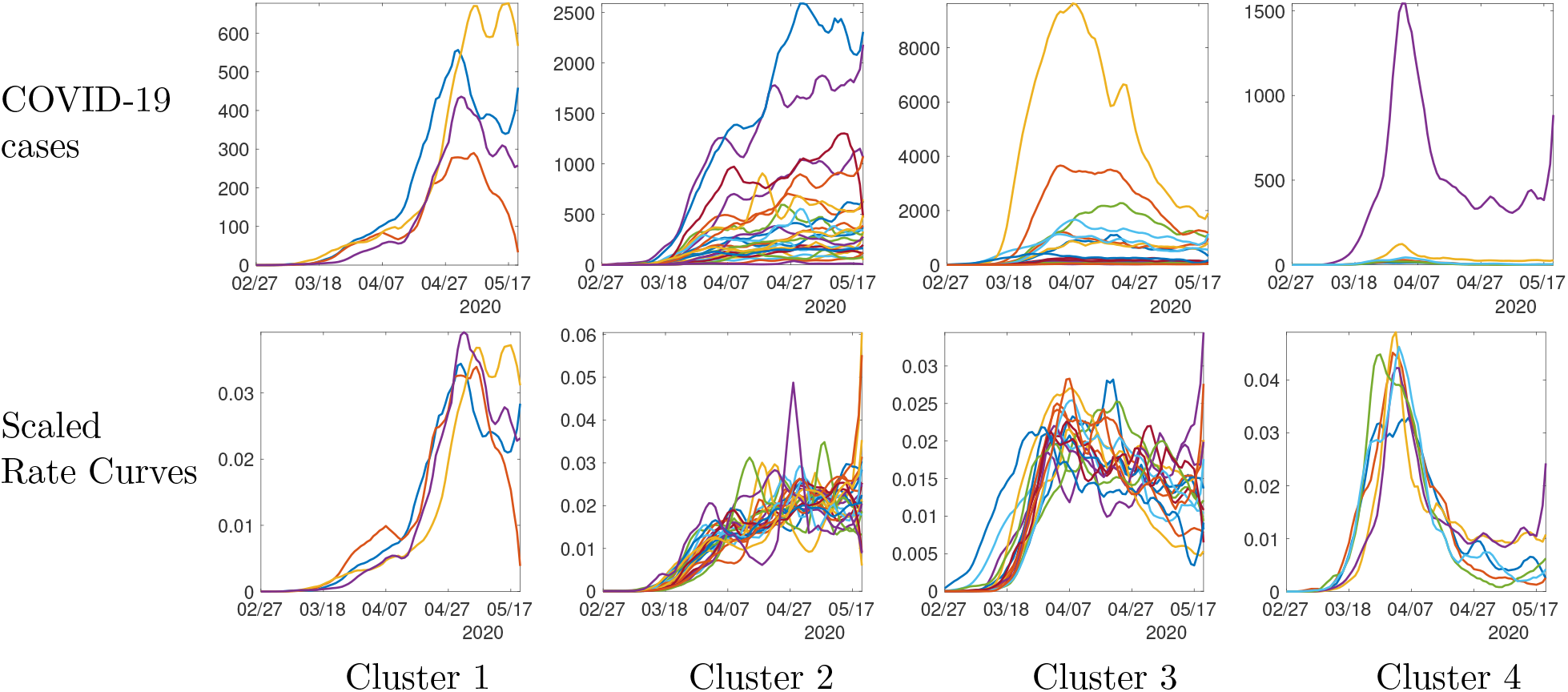
Growth rate curves for 51 US states clustered in four groups. Top row shows the curves at their original scales while the bottom row shows the normalized curves.

**Figure 5:**
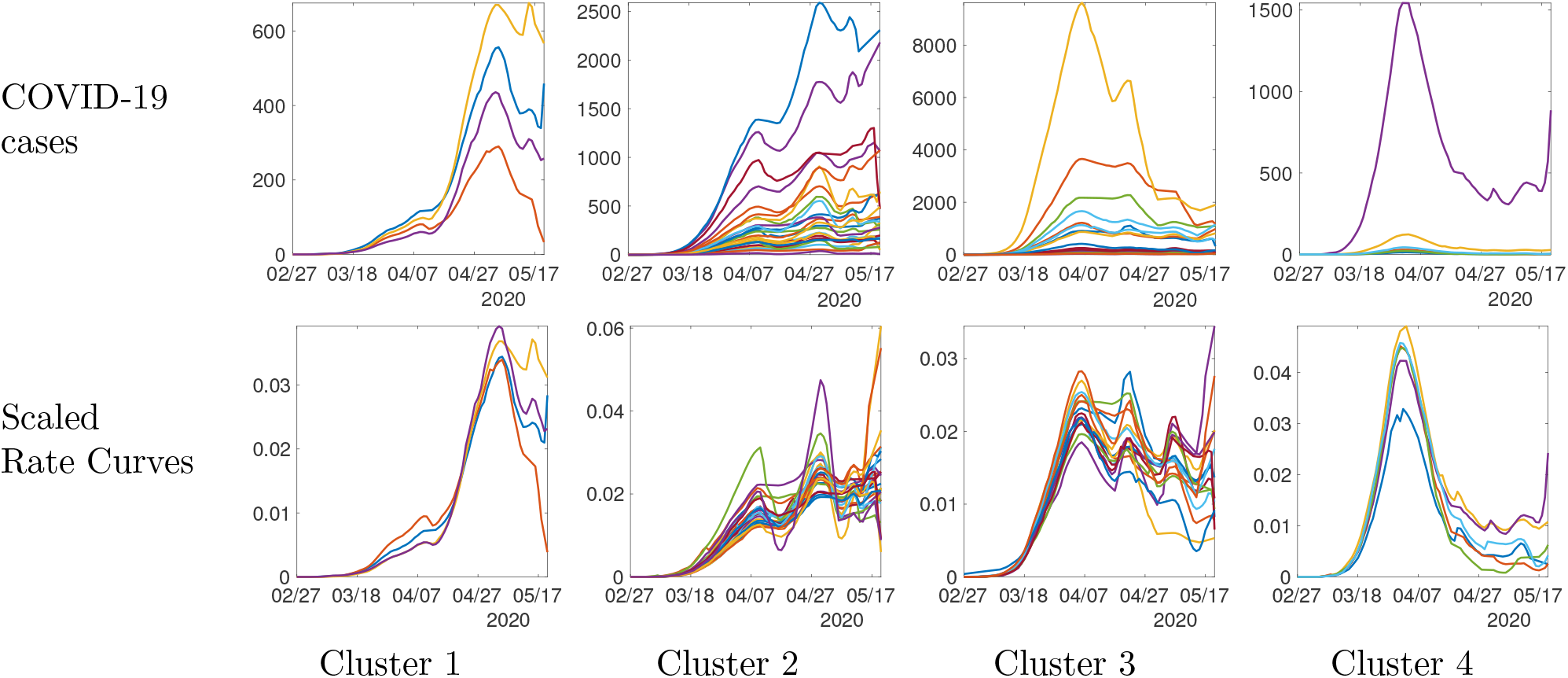
Aligned growth rate curves for 51 US states clustered in four groups. Top row shows the curves at their original scales while the bottom row shows the normalized curves.

Once the curves are aligned it is easier to visualize the common peaks and valleys (highs and lows) in each group. Figure 6 shows the average rate curves for each cluster. We obtain these curves by aligning and averaging rate curves in each cluster individually. This figure also plots a one standard-deviation band around the mean curve, in order to display the variability in the data. The pandemic growth patterns are very distinct in the four categories. For **Cluster 1:** the rate climbs rapidly and has come down only slightly; for Cluster 2, the rate climbs rapidly and then the growth slows down; for Cluster 3, the rate climbs rapidly and starts coming down slowly; and, for Cluster 4 the rate climbs rapidly and comes down all the way. The rightmost panel of this figure shows all the cluster averages in the same plot, to help visualize their differences.

**Figure 6:**
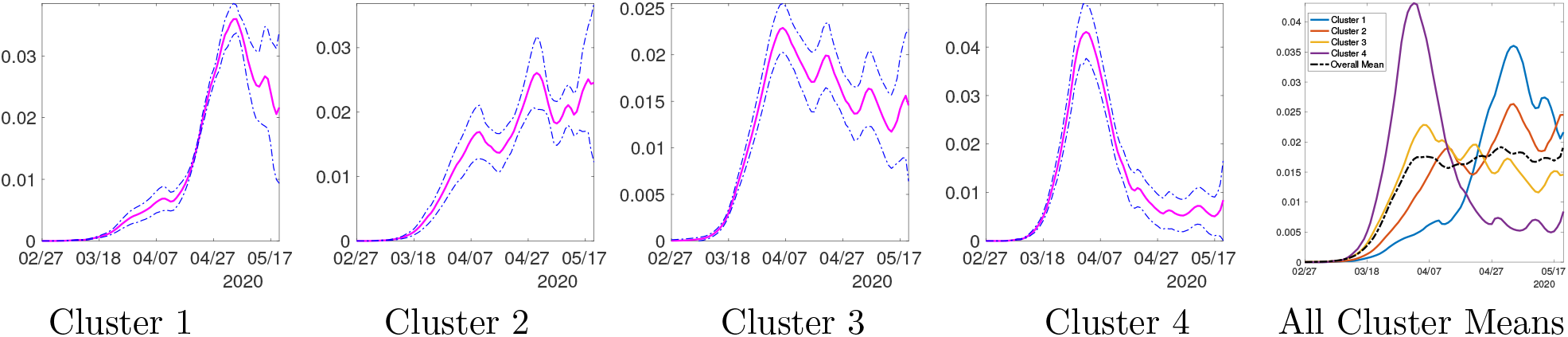
Average shapes of the growth rate curves, along with a one standard-deviation band around the mean, in each of the four clusters for the state-level USA analysis. The last panel shows all cluster averages together with the overall mean.

### 4.2 Country-level analysis in Europe

Now we present clustering and shape analysis of rate curves for 60 European countries. In this case we omit some intermediate results from clustering and alignment steps, and present the final clustering results. A dendrogram-based hierarchical clustering of countries is presented on the left side of Fig. 7. We choose to divide these countries into four clusters, as shown in the figure. The right side of the figure shows a color coding of the countries according to their cluster memberships.

**Figure 7:**
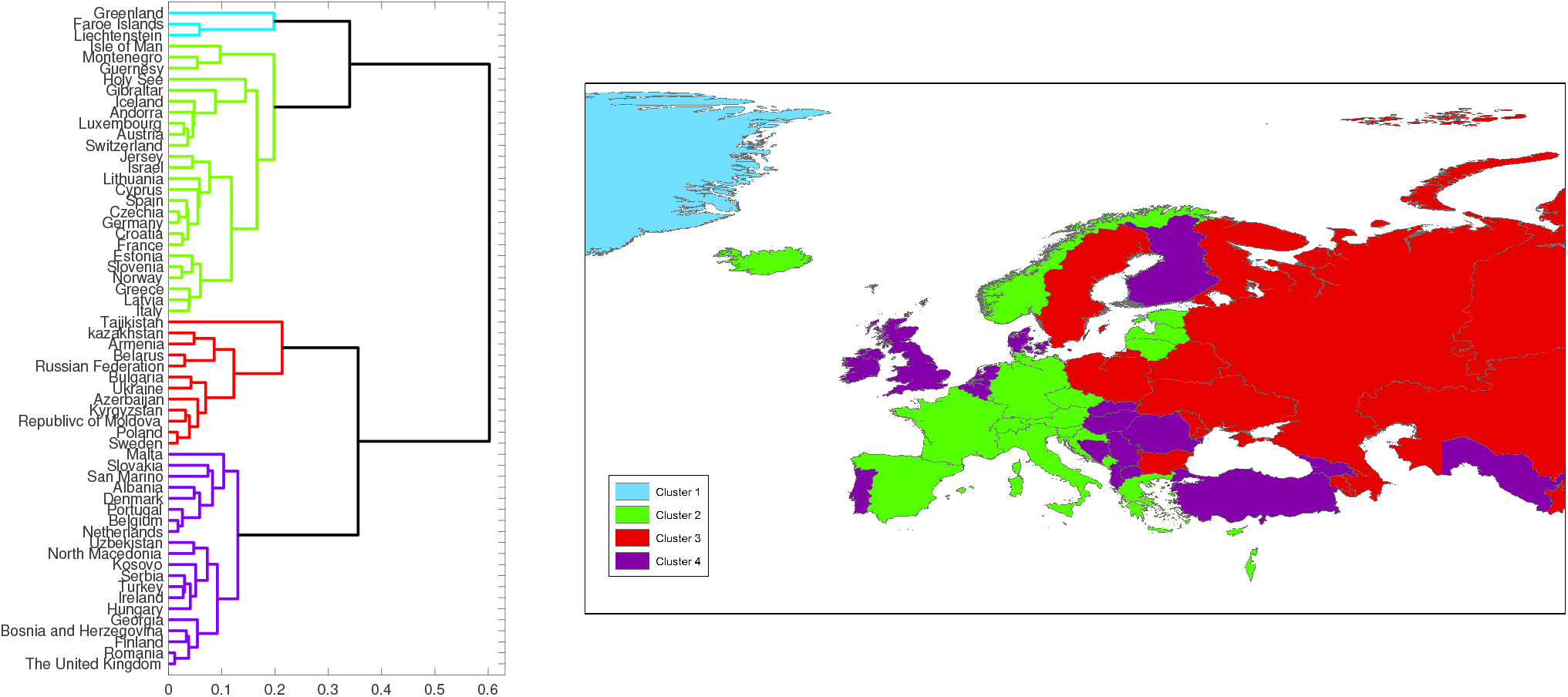
Clustering of European countries according to their normalized growth rate curves. Note that the color palette and the ordering of clusters here is different from that of USA.

The cluster membership of 60 countries is enumerated below:

- **Cluster 1:** Liechtenstein, Faroe Islands, Greenland
- **Cluster 2:** Italy, France, Germany, Spain, Switzerland, Norway, Austria, Croatia, Israel, Czechia, Greece, Iceland, Andorra, Estonia, Holy See, Latvia, Luxembourg, Lithuania, Slovenia, Gibraltar, Guernesy, Cyprus, Jersey, Montenegro, Isle of Man.
- **Cluster 3:** The United Kingdom, Netherlands, Denmark, Georgia, Romania, Finland, Portugal, Belgium, Bosnia and Herzegovina, Hungary, Ireland, North Macedonia, San Marino, Serbia, Slovakia, Malta, Albania, Turkey, Uzbekistan, Kosovo.
- **Cluster 4:** Sweden, Azerbaijan, Russian Federation, Belarus, Armenia, Poland, Ukraine, Republic of Moldova, Kazakhstan, Kyrgyzstan, Tajikistan.

Once the curves are aligned within their clusters, it is easier to visualize the common peaks and valleys (highs and lows) in each group. Figure 8 shows the average normalized growth curves for each cluster. We obtain these curves by aligning and averaging normalized curves in each cluster individually. The growth patterns are very distinct in the four categories. For **cluster 1**, the rate climbs rapidly and comes down all the way to the normal levels. For **cluster 2**, the rate climbs rapidly and starts coming down slowly, while in **cluster 3**, the rate has only started to come down slowly. In case of **cluster 4**, the growth rate is still climbing rapidly. The rightmost panel shows all the cluster averages in the same plot.

**Figure 8:**
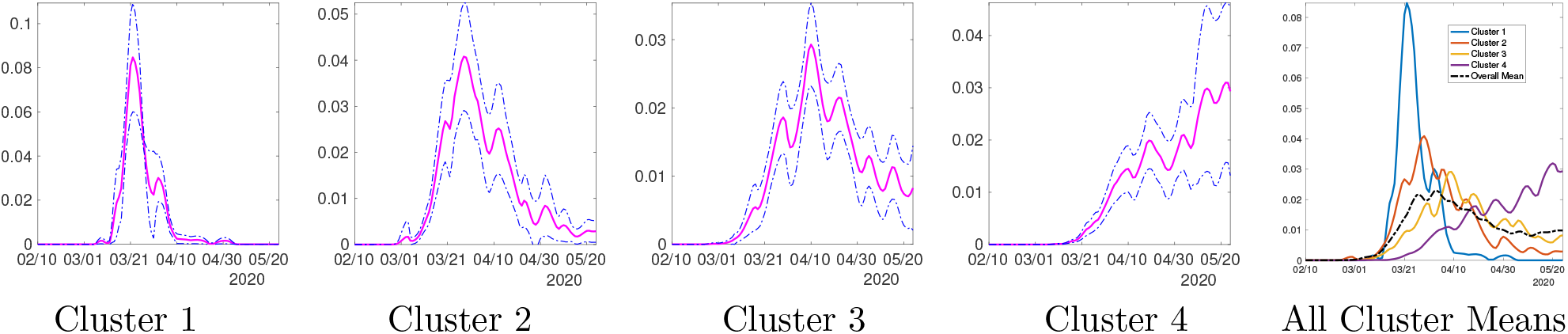
Average shapes of the growth rate curves in each of the four clusters in Europe. The last panel shows all averages together with the overall mean.

## 5 Discussion

In this paper we employ well-established methods for classifying and synthesizing *shapes* of growth rate curves underlying the spatial heterogeneity that exists at finer spatial scales. Specifically, we have applied this methodology to characterize the spatial-temporal dynamics of the pandemic at two different scales of spatial aggregation: Across states within the USA and across nations withi7n Europe. Our analyses reveal a few dominant incidence trajectories that characterize transmission dynamics at the state level in the USA and at the country level in Europe. Overall, our approach reveals broad classifications of spatial areas into different trajectories and adds to the methodological toolkit for guiding public health decision making during epidemic emergencies at different spatial scales.

The main findings are:

- The shapes of rates curves (or incidence curves) are different across the spatial units, but they cluster into a few groups with characteristic patterns.
- The resulting broad classifications of the shapes (e.g., rapidly rising, slowly rising, slowly decreasing, and declining and reaching low incidence levels) are clearly visible and very meaningful.
- Computing average growth rates within the clusters is more appropriate than taking raw averages across all spatial areas comprising the population, particularly when the epidemic is comprised by asynchronous outbreaks.

Our state-level analysis indicate that Alaska, Hawaii, Idaho, Louisiana, Montana, and Vermont are the only states that appear to be bringing the COVID-19 epidemic under control (Cluster 4). The other three characteristic patterns that emerge from our analysis paint a grim picture of the COVID-19 epidemic in the USA at a time when most of the states have begun to reopen their economies at least in some way. In comparison, our country-level analysis of the epidemic in Europe reveal that Liechtenstein, Faroe Islands, and Greenland appear to have brought their epidemics under control while countries in Cluster 2 are characterized by a downward trend and appear to be in process of controlling their epidemics. In contrast, the epidemic is following an alarming increasing trend in Sweden, Azerbaijan, Russian Federation, Belarus, Armenia, Poland, Ukraine, Republic of Moldova, Kazakhstan, Kyrgyzstan, and Tajikistan while several other countries (Cluster 3) appear to be reaching an endemic level of the disease.

It is worth noting that the methodology employed here requires little human intervention. The only quantity that may be specified manually is the number of clusters, and that is left as a choice for the end user. Hence, the clusters of spatial units that emerge from the analysis provides an objective classification system of epidemic patterns, which can be used to guide the implementation or relaxation of public health measures as the epidemic emergency evolves over time. For instance, the resulting configuration of clusters for the state-level analysis in the US states using a truncated dataset for an earlier period *02/27/20 - 04/25/20* is presented in Fig. 9 where the average rate curves of the four clusters are displayed. In the bottom panel we show the clustering labels for 50 states. Comparing this map with that obtained from the analysis using the entire dataset (Fig. 3), we see that a number of states have changed cluster memberships while many others remain in the same cluster. States along the northeastern, western, and southeastern borders have shifted to the next worst cluster in terms of the higher growth rates.

**Figure 9:**
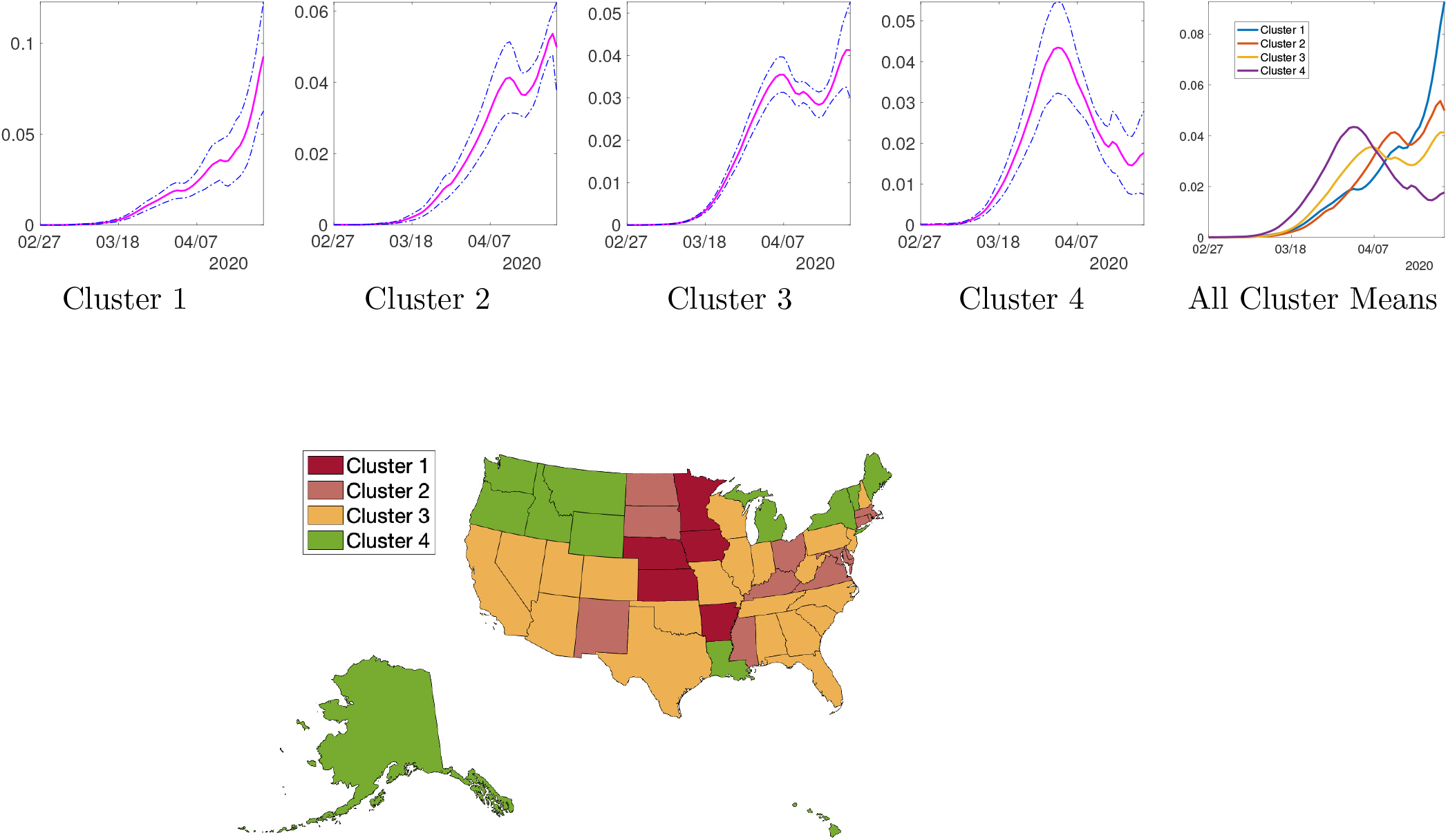
Top: Average rate curves for four clusters resulting from the data collected up to April 25th. Bottom: The corresponding clustering of 50 US states.

While we have focused our analysis on the time series of confirmed cases across spatial areas, our analysis could be extended to consider the trajectory of the epidemic in terms of the number of reported COVID-19 deaths. Moreover, a limitation of our analysis stems from the fact that testing rates have generally improved across spatial areas during the course of the pandemic, which could have influenced the shapes of the epidemic curves particularly during the early transmission phase. In particular, the process of ramping up testing rates took several weeks in the USA, and accumulating evidence suggests that the epidemic in the USA and Europe likely started much earlier than initially thought.

## Data Availability

The data used here is publicly available at websites cited in the article.

https://www.who.int/emergencies/diseases/novel-coronavirus-2019/events-as-they-happen

## Acknowledgement

This research was supported in part by the grants NIH R01 GM135927 and NIH R01 MH120299 to AS and grants NSF 1414374 as part of the joint NSF-National Institutes of Health NIH-United States Department of Agriculture USDA Ecology and Evolution of Infectious Diseases program; UK Biotechnology and Biological Sciences Research Council [grant BB/M008894/1] and RAPID NSF 2026797 to GC.

## Notes

### Competing Interest Statement

The authors have declared no competing interest.

### Author Declarations

There was no experimentation involved in this research.

